# The impact of climate temperature on counts, recovery, and death rates due to SARS-CoV-2 in South Africa

**DOI:** 10.1101/2020.07.16.20155523

**Authors:** Naven Chetty, Bamise Adeleye, Abiola Olawale Ilori

## Abstract

The impact of climate temperature on the counts (number of positive COVID-19 cases reported), recovery, and death rates of COVID-19 cases in South Africa’s nine provinces was investigated. The data for confirmed cases of COVID-19 were collected for March 25 and June 30, 2020 (14 weeks) from South Africa’s Government COVID-19 online resource, while the daily provincial climate temperatures were collected from the website of the South African Weather Service. Our result indicates that a higher or lower climate temperature does not prevent or delay the spread and death rates but shows significant positive impacts on the recovery rates of COVID-19 patients. Thus, it indicates that the climate temperature is unlikely to impose a strict limit on the spread of COVID-19. There is no correlation between the cases and death rates, an indicator that no particular temperature range is closely associated with a faster or slower death rate of COVID-19 patients. As evidence from our study, a warm climate temperature can only increase the recovery rate of COVID-19 patients, ultimately impacting the death and active case rates and freeing up resources quicker to enable health facilities to deal with those patients’ climbing rates who need treatment.

## 1. Introduction

The roles of climate temperature on transmitting the novel Severe Acute Respiratory Syndrome Coronavirus (SARS-CoV-2) and patients’ recovery and death rates seemed to have generated significant worldwide debates. Several kinds of research indicated the spreads of other seasonal flu virus outbreaks by reporting a substantial number of cases with colder climate conditions similar to the spreads of SARS-CoV-2 [1, 2]. In contrast, others found an inverse or no correlation between climate temperature variations and the Coronavirus [3–5].

SARS-CoV-2, popularly called Coronavirus (COVID-19), envelopes positive-stranded RNA viruses with a characteristic surface [6, 7]. It has been reported that the infection of the virus in humans causes severe gastroenteritis and respiratory tract diseases [6, 8]. These viruses’ persistence was officially lost at around 56°C but somewhat differed depending on the infection [9]. In their study of SARS-CoV-2 aerosol and surface stability, Doremalen *et al*. [10] revealed that the virus could survive on hard surfaces for up to 72 hours at temperatures between 21°C and 23°C. Similarly, Zhou *et al*. [11] found the coronavirus to be most active between 9°C and 24°C, decreasing with higher temperatures. The fact that the virus causing the SARS-CoV-2 is new still limits information about how it impacted by variations in climatic temperature conditions. Peng *et al*. [2] studied the impacts of temperature and absolute humidity on COVID-19 in 31 provincial regions of mainland China. They observed a decline in the transmission rate of the COVID-19 as temperature increases. A recent study examining the impact of temperature variation and humidity on COVID-19 mortality in Wuhan China found that mortality rates were lower for days with higher temperatures and humidity. The study also indicates high death rates on days with a broader maximum and minimum temperature range [12].

Therefore, this study aims to investigate the impact of climate temperature variation on the counts, recovery, and death rates of COVID-19 cases in all South Africa’s provinces. The findings were compared with those of countries with comparable climate temperature values.

## 2. Methods

### 2.1 Data Collection

The data for confirmed cases of COVID-19 were collected for March 25 and June 30 (14 weeks) for South African provinces, including daily counts, death, and recovery rates. The dates were grouped into two, wherein weeks 1-5 represent the periods of total lockdown to contain the spread of COVID-19 in South Africa. Weeks 6-14 are periods where the lockdown was eased to various levels 4 and 3. The daily information of COVID-19 count, death, and recovery was obtained from South Africa’s Government COVID-19 online resource (https://sacoronavirus.co.za). Daily provincial climate temperatures were collected from the website of the South African Weather Service (https://www.weathersa.co.za). The provinces of South Africa are Eastern Cape, Western Cape, Northern Cape, Limpopo, Northwest, Mpumalanga, Free State, KwaZulu-Natal, Western Cape, and Gauteng. Weekly consideration was given to the daily climate temperature (average minimum and maximum). The recorded values were considered, respectively, to be in the ratio of death-to-count (D/C) and recovery-to-count (R/C).

### 2.2 Statistical Analysis

Descriptive statistics were performed for all the data collected for this study. The analyses were performed using the Person’s bivariate correlation to analyze the association between climate temperature, death-to-count, and recovery-to-count ratios of COVID-19. The death and recovery rate were first accommodated to check the ratio of weekly COVID-19 counts to death/recovery rates. Then, the climate temperature was introduced into the equation to verify the correlation between the parameters of interests. Equations 1 and 2 used are given as a pair of variables (C, D/C, and R/C) [14-16].

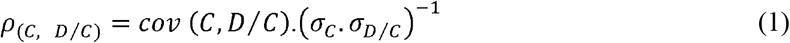

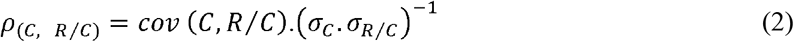

Where *ρ*_(*C, D/C*)_ is the correlation between counts and death-to-count ratio; *ρ*_(*C, R/C*)_ is the correlation between counts and recovery-to-count ratio; *cov* is the covariance of the count and the interested ratios; *σ*_*C*_, *σ*_*D/C*,_ *σ*_*R/C*_ are the standard deviation of *C, D*/*C*, and *R*/*C* respectively. The correlation curves between the parameters were fitted to get the coefficient of determination (R^2^). The analysis was performed using OriginPro 8.5.0 SR1.

## 3. Results and Discussion

### 3.1 Description of COVID-19 counts, death and recovery rates within the study period

The average climate temperature within the study period for Eastern Cape Province ranges from 10.2 to 29.6 °C with 0.0295 (2.95 %) death-to-count ratio and 0.0207 (2.07 %) recovery-to-count ratio for the lockdown periods and average climate temperature of 6.8 to 29.4 °C with 0.0204 (2.04 %) death-to-count ratio and 0.0683 (6.83 %) recovery-to-count ratio for the eased-lockdown periods, respectively. The average climate temperature within the study period for Northern Cape Province ranges from 9.2 to 33.8^°^C with 0.1000 (10 %) death-to-count ratio and 1.400 (140 %) recovery-to-count ratio for the lockdown periods and average climate temperature of 1.7 to 28.3 °C with 0.0000 (0 %) death-to-count ratio and 0.5732 (57.32 %) recovery-to-count ratio for the eased-lockdown periods, respectively. The average climate temperature within the study period for Limpopo Province ranges from 10.8 to 29.8 °C with 0.1833 (18.33 %) death-to-count ratio and 1.1667 (116.67 %) recovery-to-count ratio for the lockdown periods and average climate temperature of 4.4 to 25.2 °C with 0.0095 (0.95 %) death-to-count ratio and 0.4794 (47.94 %) recovery-to-count ratio for the eased-lockdown periods, respectively. The average climate temperature within the study period for Northwest Province ranges from 9.4 to 30 °C with 0.000 (0 %) death-to-count ratio and 0.8412 (84.12 %) recovery-to-count ratio for the lockdown periods and average climate temperature of 0.5 to 25.1 °C with 0.0073 (0.73 %) death-to-count ratio and 0.1889 (18.89 %) recovery-to-count ratio for the eased-lockdown periods, respectively. The average climate temperature within the study period for Mpumalanga Province ranges from 10.4 to 30 °C with 0.000 (0 %) death-to-count ratio and 0.8400 (84 %) recovery-to-count ratio for the lockdown periods and average climate temperature of 7.1 to 25.9 °C with 0.0033 (0.33 %) death-to-count ratio and 0.5681 (56.81 %) recovery-to-count ratio for the eased-lockdown periods, respectively. The average climate temperature within the study period for Free State Province ranges from 4.8 to 28.4 °C with 0.1071 (10.71 %) death-to-count ratio and 2.0232 (202.32 %) recovery-to-count ratio for the lockdown periods and average climate temperature of -2.7 to 24 °C with 0.0098 (0.98 %) death-to-count ratio and 0.3362 (33.62 %) recovery-to-count ratio for the eased-lockdown periods, respectively. The average climate temperature within the study period for KwaZulu-Natal Province ranges from 15.6 to 29.6 °C with 0.0388 (3.88 %) death-to-count ratio and 0.3342 (33.42 %) recovery-to-count ratio for the lockdown periods and average climate temperature of 10.4 to 28.7 °C with 0.0146 (1.46 %) death-to-count ratio and 0.5626 (56.26 %) recovery-to-count ratio for the eased-lockdown periods, respectively. The average climate temperature within the study period for Western Cape Province ranges from 7.6 to 28.6 °C with 0.0188 (1.88 %) death-to-count ratio and 0.2933 (29.33 %) recovery-to-count ratio for the lockdown periods and average climate temperature of 6.7 to 25 °C with 0.0274 (2.74 %) death-to-count ratio and 0.5967 (59.67 %) recovery-to-count ratio for the eased-lockdown periods, respectively. The average climate temperature within the study period for Gauteng Province ranges from 8.8 to 24.6 °C with 0.0066 (0.66 %) death-to-count ratio and 0.8233 (82.33 %) recovery-to-count ratio for the lockdown periods and average climate temperature of 1.6 to 20.6 °C with 0.0099 (0.99 %) death-to-count ratio and 0.4077 (40.77 %) recovery-to-count ratio for the eased-lockdown periods, respectively.

The results of COVID-19 cases, death, recovery rates along with the death-to-count, recovery-to-count ratios, and average weekly climate temperature are presented in Table 1 (a-i).

**Table 1 (a – i):**
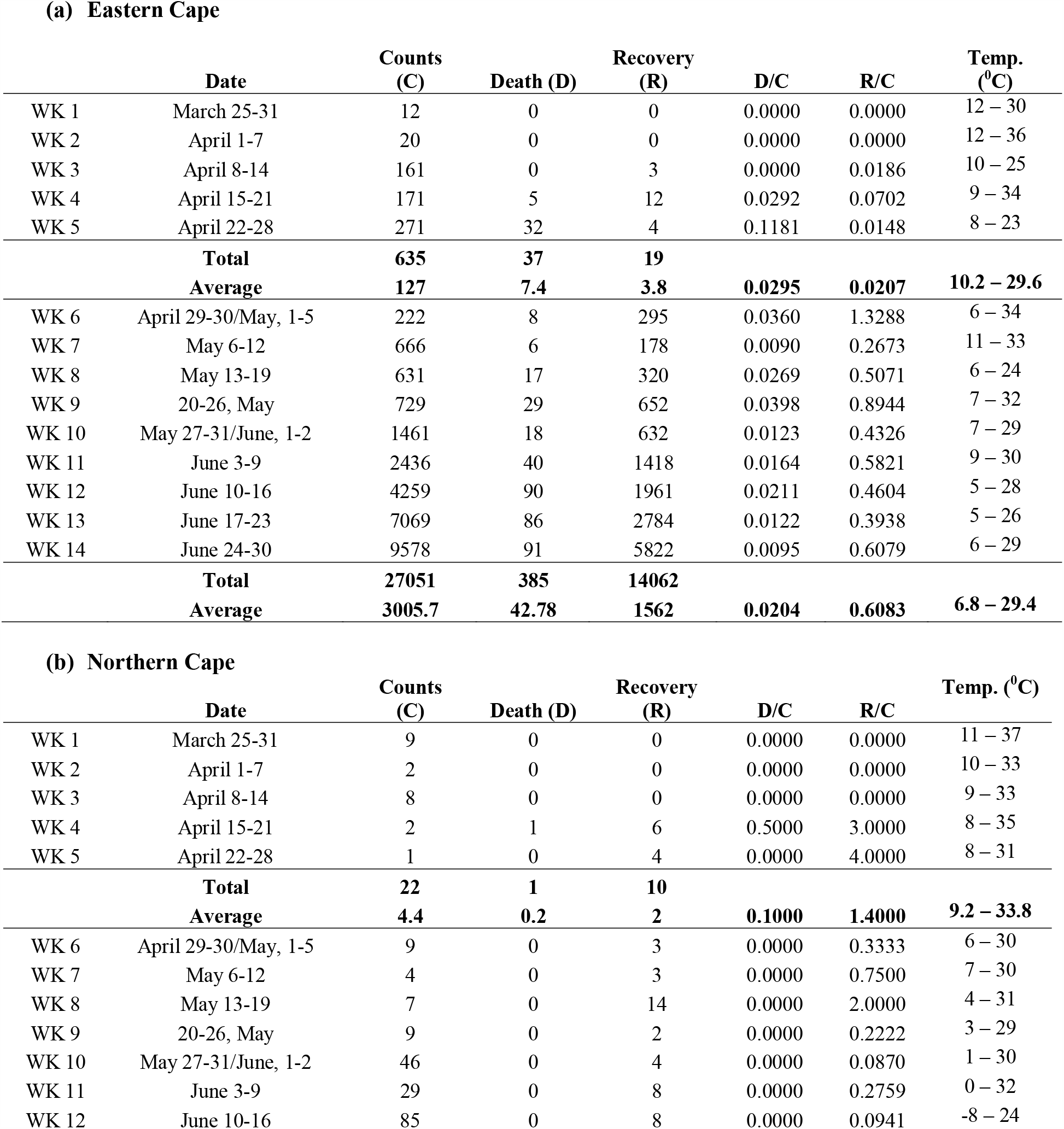
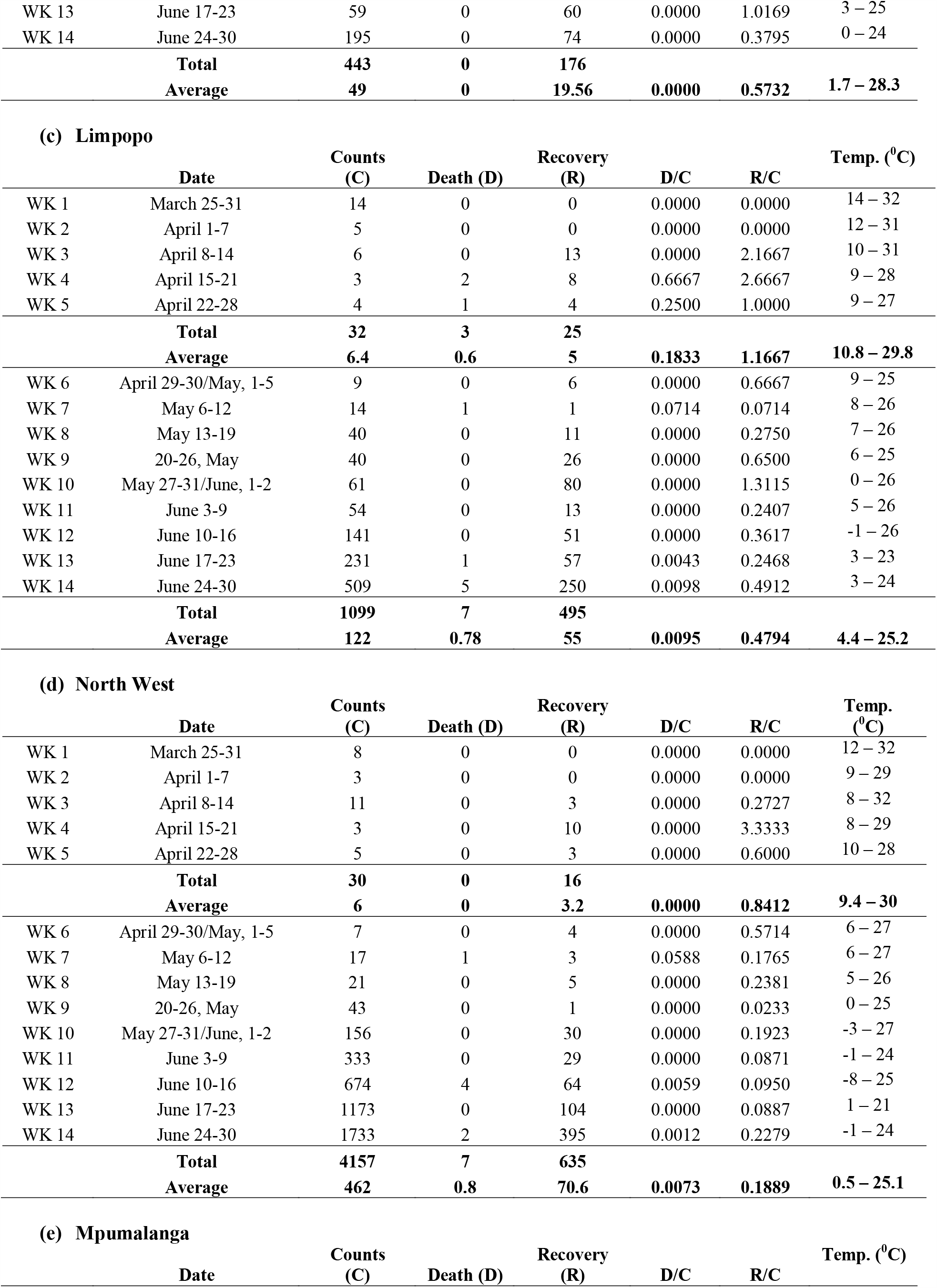
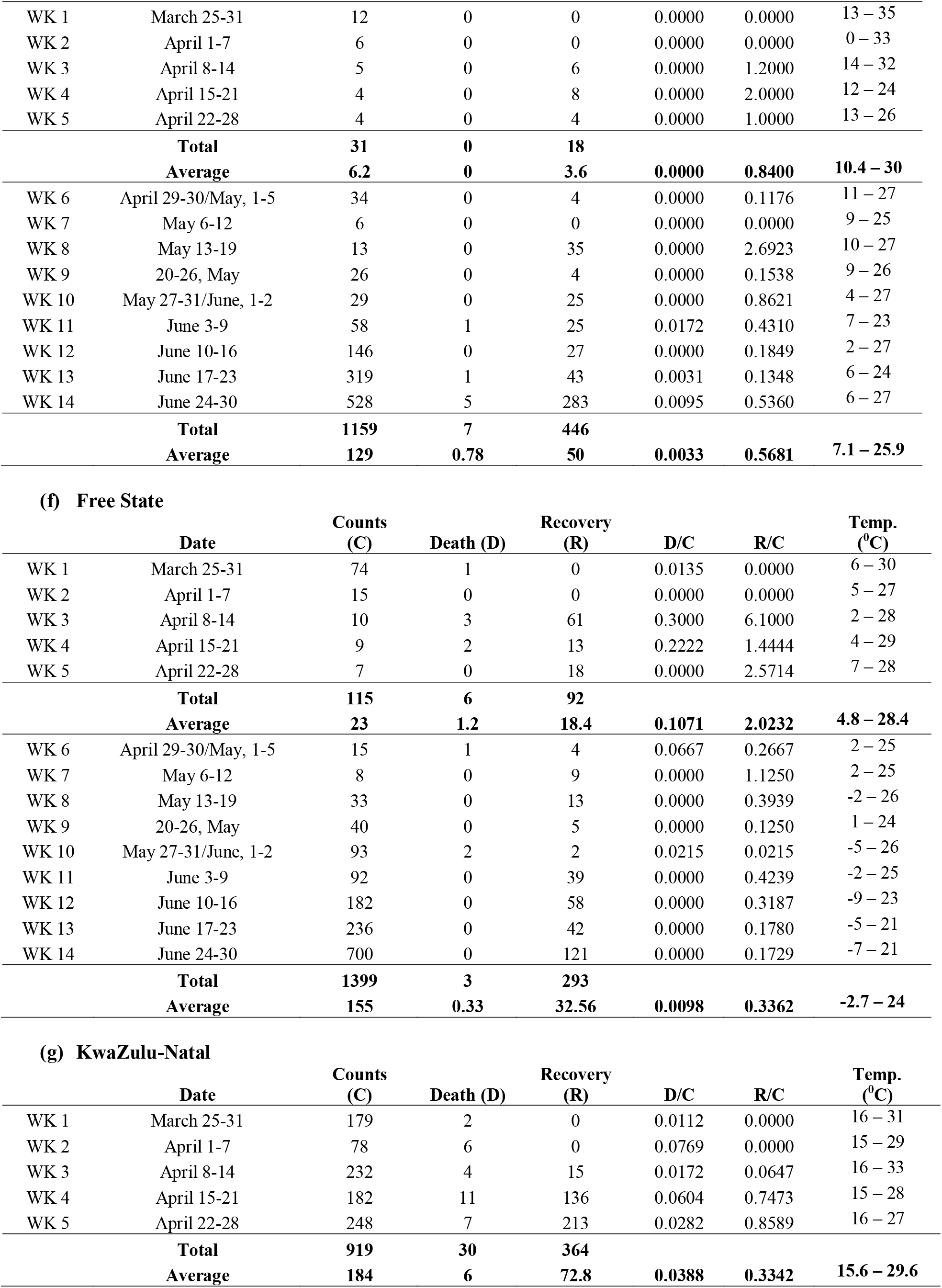
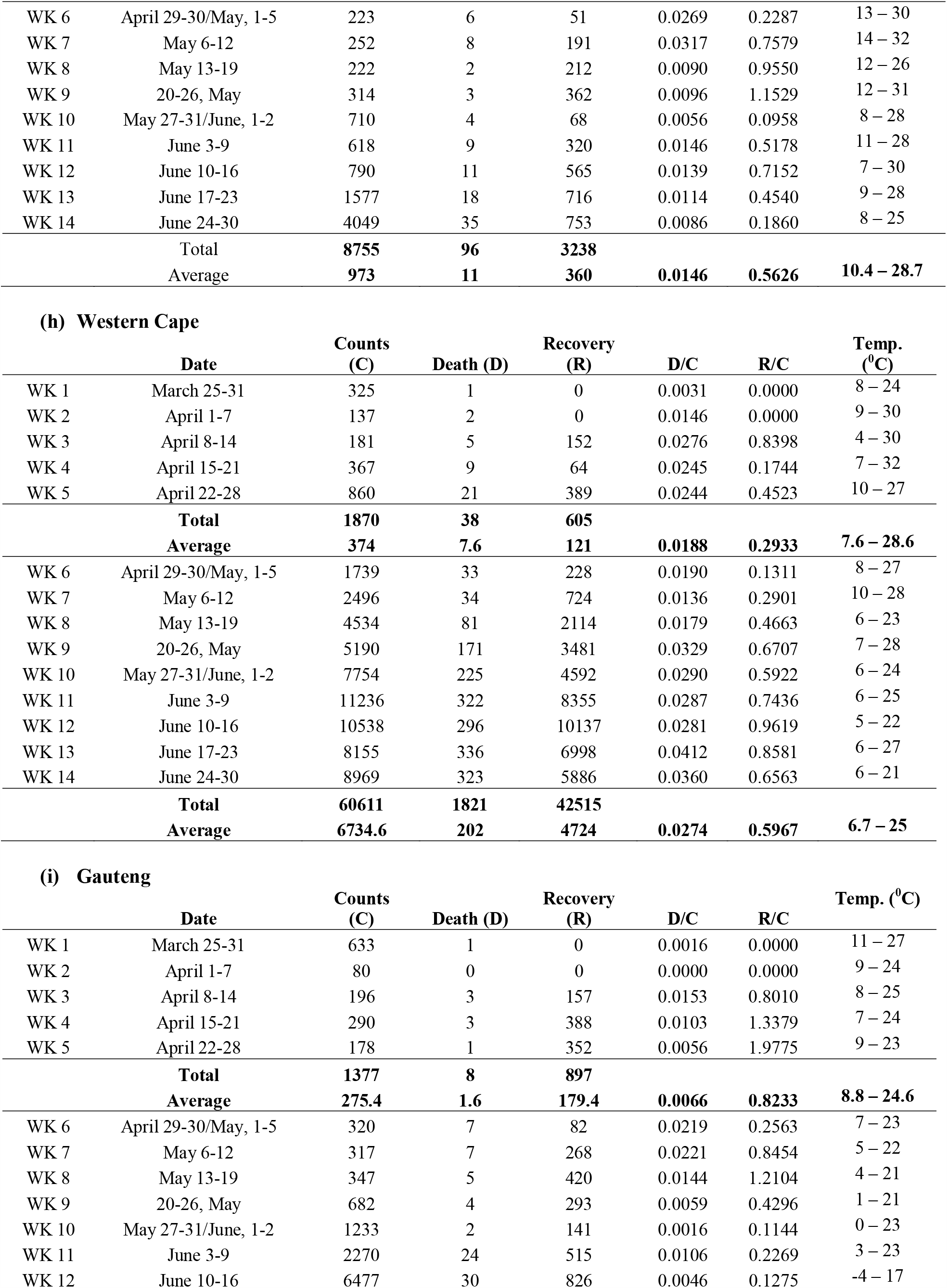
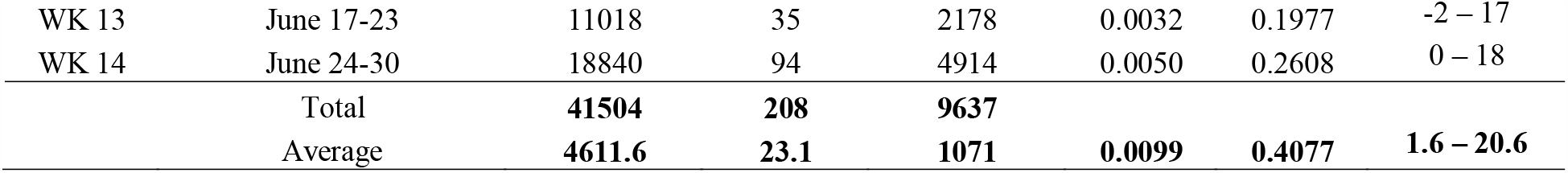
COVID-19 counts, death, recovery rates along with the death-to-count, recovery-to-count ratios, and average weekly climate temperature.

As shown from the results in Table 1 (a – i), higher or lower climate temperatures cannot be reported to prevent or delay the transmissions and death rates of patients of COVID-19. Nonetheless, it can be concluded that a higher temperature of the climate raises recovery rates for COVID-19 patients. Figure 2 shows the average minimum and maximum climate temperatures and the death-to-count ratio for the lockdown period. In contrast, Figure 3 shows the average minimum and maximum climate temperatures and the COVID-19 recovery-to-count ratio for the lockdown period.

**Fig 1:**
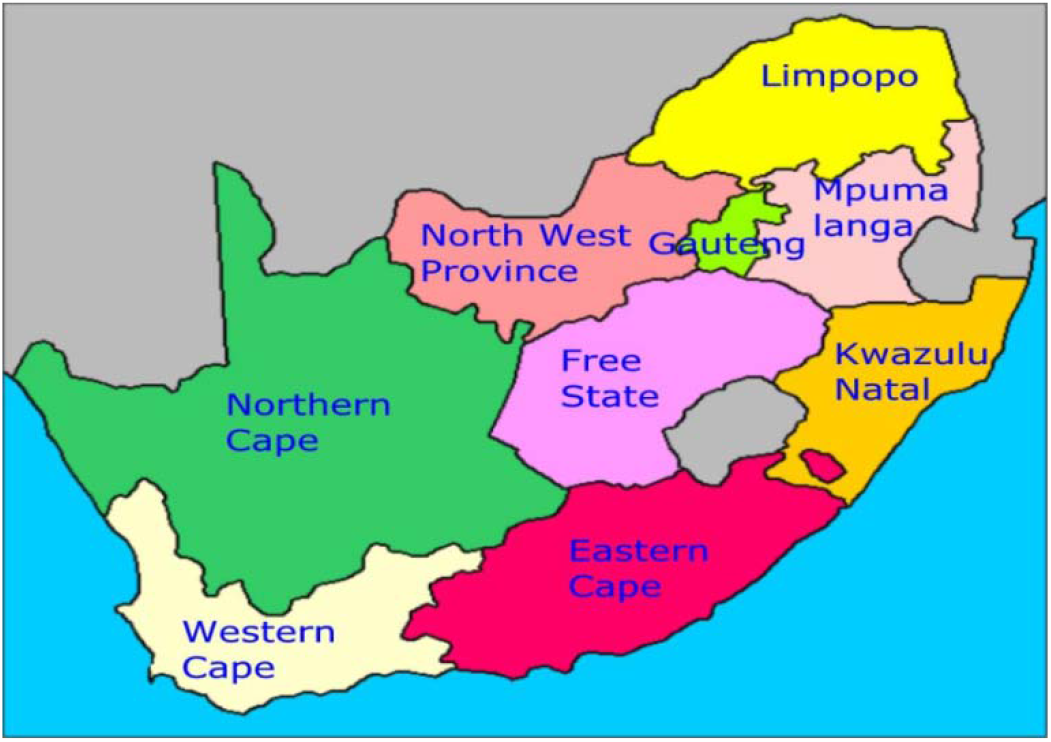
Map of South Africa showing the provinces [13].

**Fig 2:**
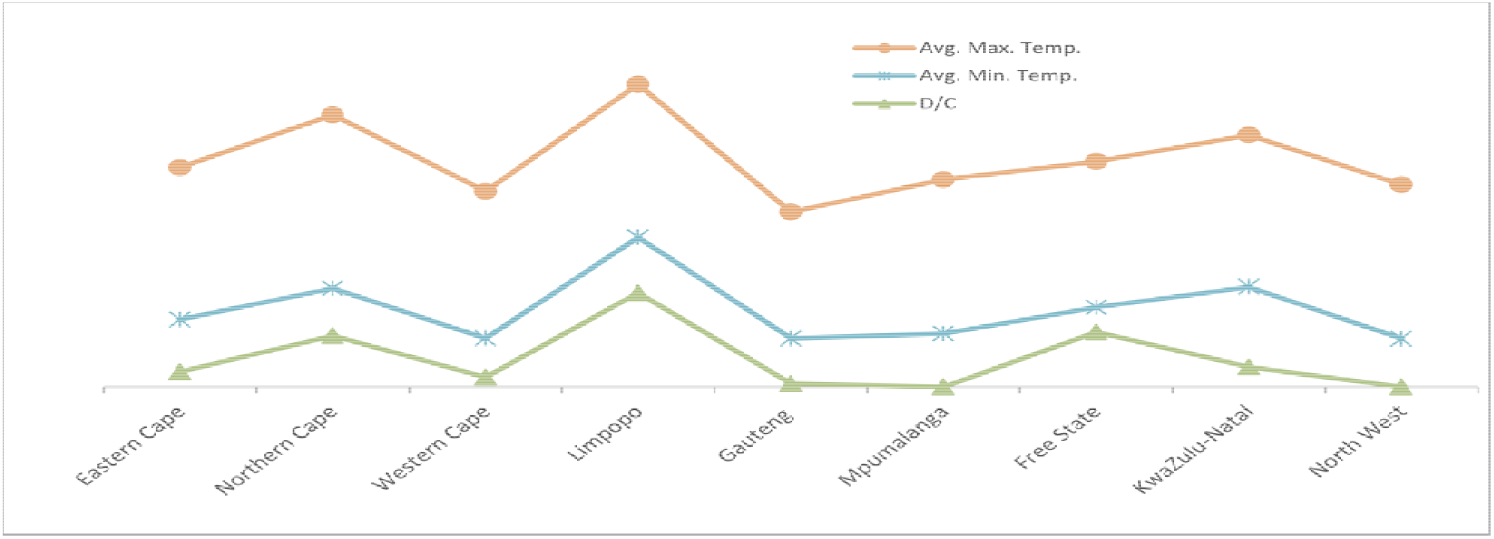
Average minimum and maximum climate temperatures and death-to-count (D/C) ratio for the lockdown period.

**Fig 3:**
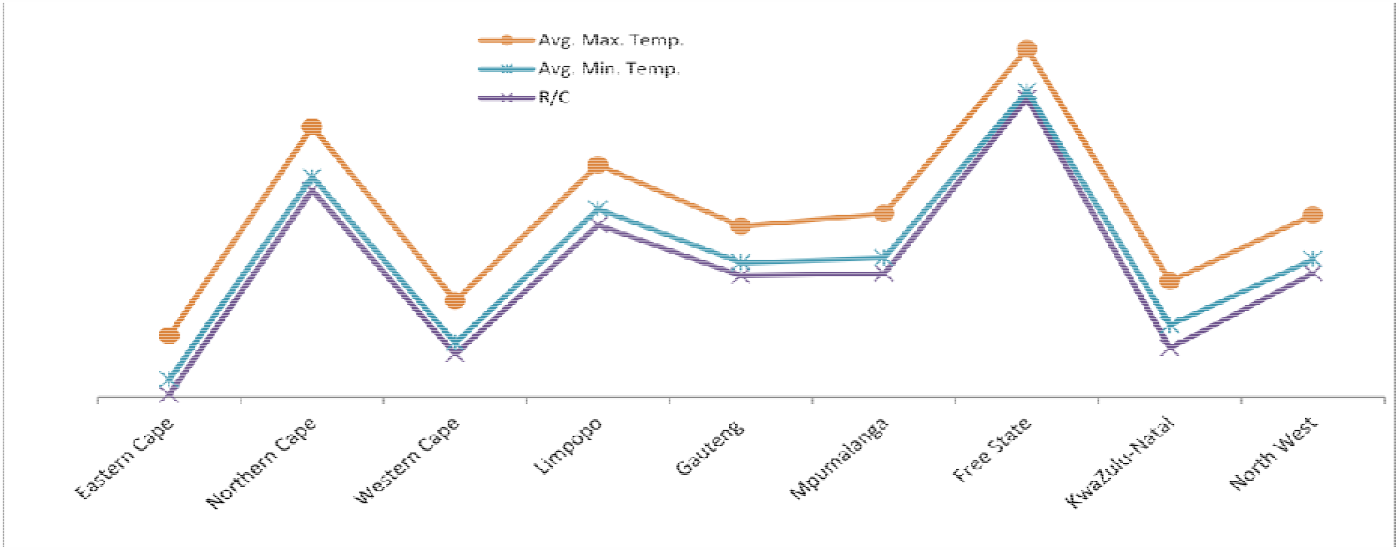
Average minimum and maximum climate temperatures and recovery-to-count (R/C) for the lockdown period.

Figure 4 shows the average minimum and maximum climate temperatures and the death-to-count ratio for the eased-lockdown period. In contrast, Figure 5 shows the average minimum and maximum climate temperatures and the COVID-19 recovery-to-count ratio for the eased-lockdown period.

**Fig 4:**
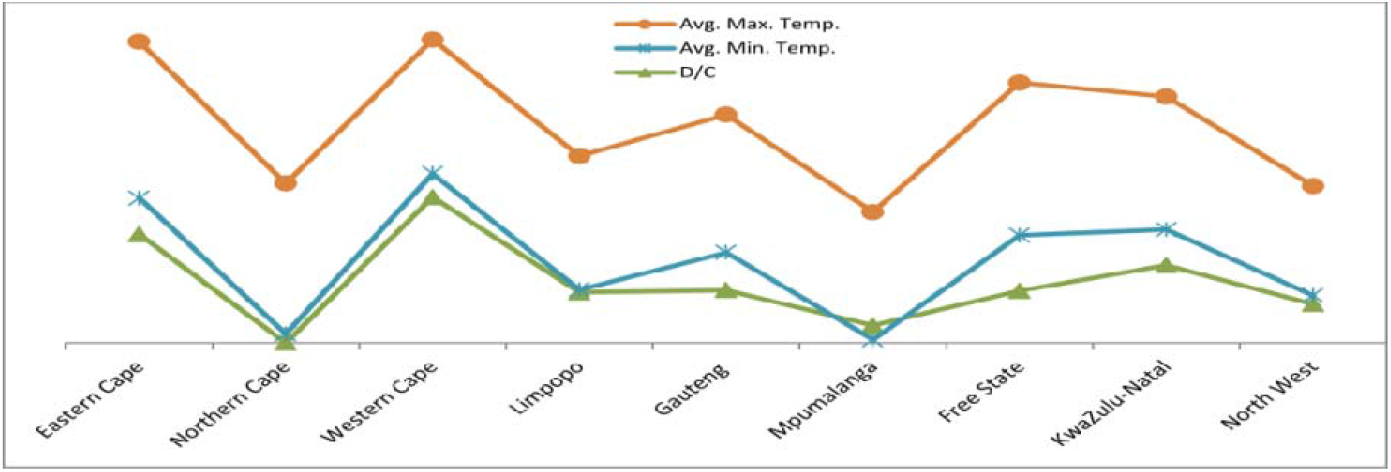
Average minimum and maximum climate temperatures and death-to-count (D/C) ratio for the eased-lockdown period.

**Fig 5:**
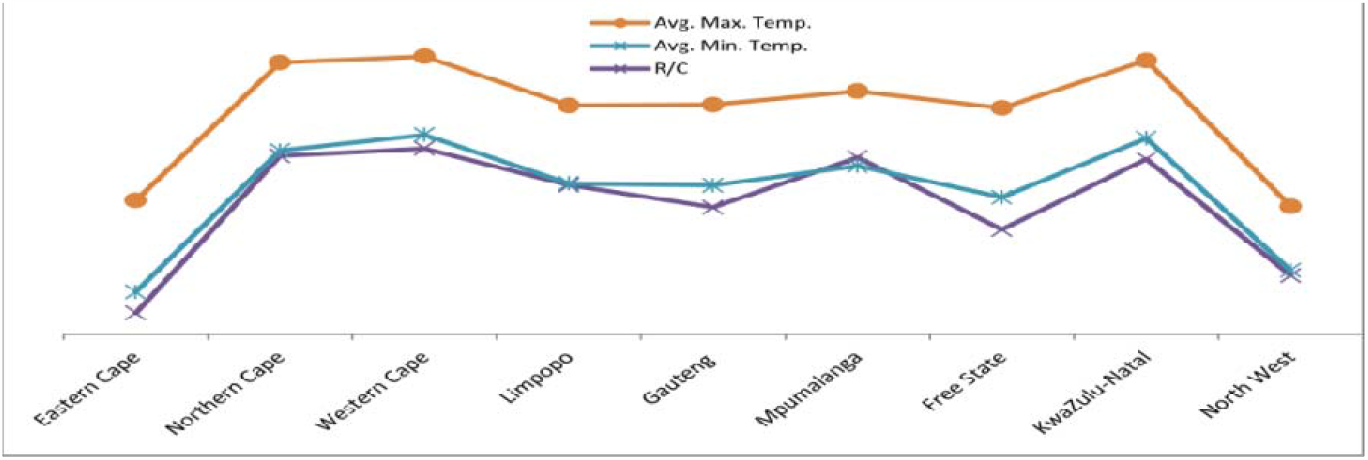
Average minimum and maximum climate temperatures and recovery-to-count (R/C) for the eased-lockdown period.

The results showed that higher climate temperatures aren’t essential to avoid the COVID-19 from being spread. The present results conform to the reports that suggested that COVID-19 is unlike the seasonal flu, which does dissipate as the climate temperature rises [17]. Accordingly, the ratio of counts and death-to-count cannot be concluded to be influenced by variations in the climate temperatures within the study areas.

### 3.2 Correlation between climate temperature, death-to-count and recovery-to-death ratio of COVID-19 in the study areas

Table 2 showed the correlation values for climate temperatures, the death-to-count, and the recovery-to-count ratios of COVID-19 in South African provinces during the study period (March 25 to June 30, 2020). Figures 6 and 7 presented the correlation pattern of COVID-19 death and recovery rates and the daily minimum and maximum temperatures in the study period.

**Table 2:**
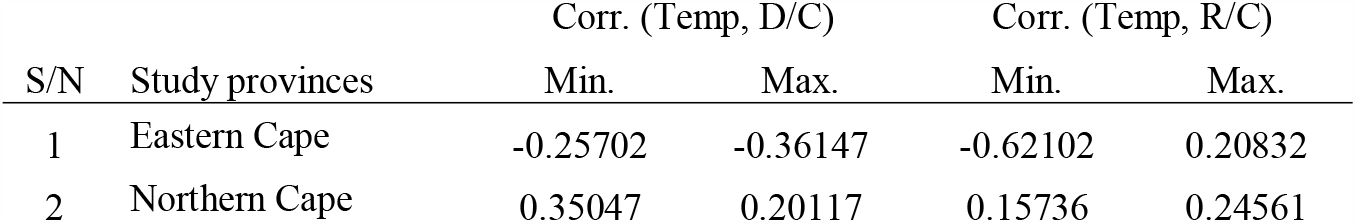
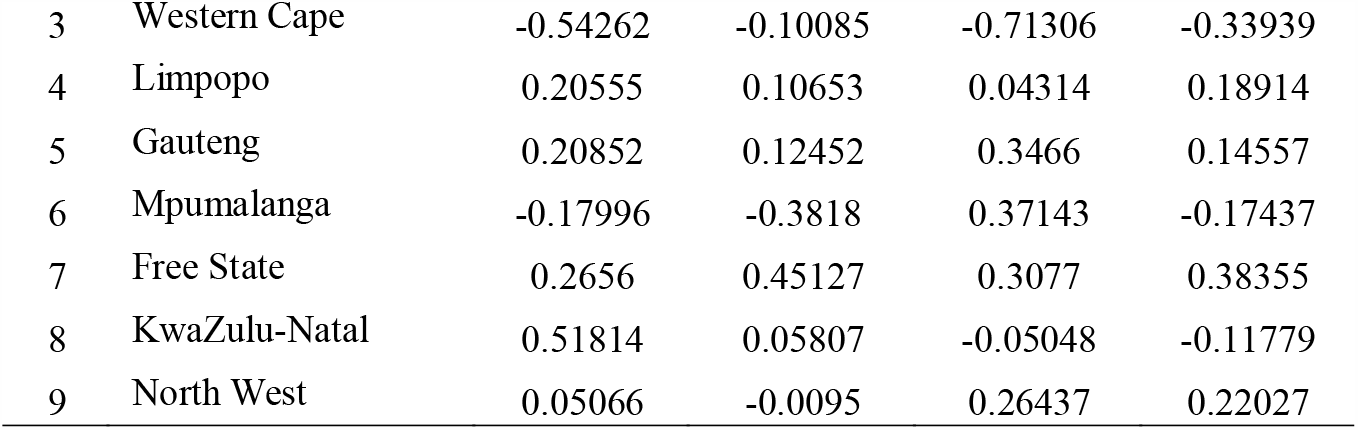
Correlation of climate temperature with death-to-count, and recovery-to-count rates of COVID-19 within the study period.

**Figure 6:**
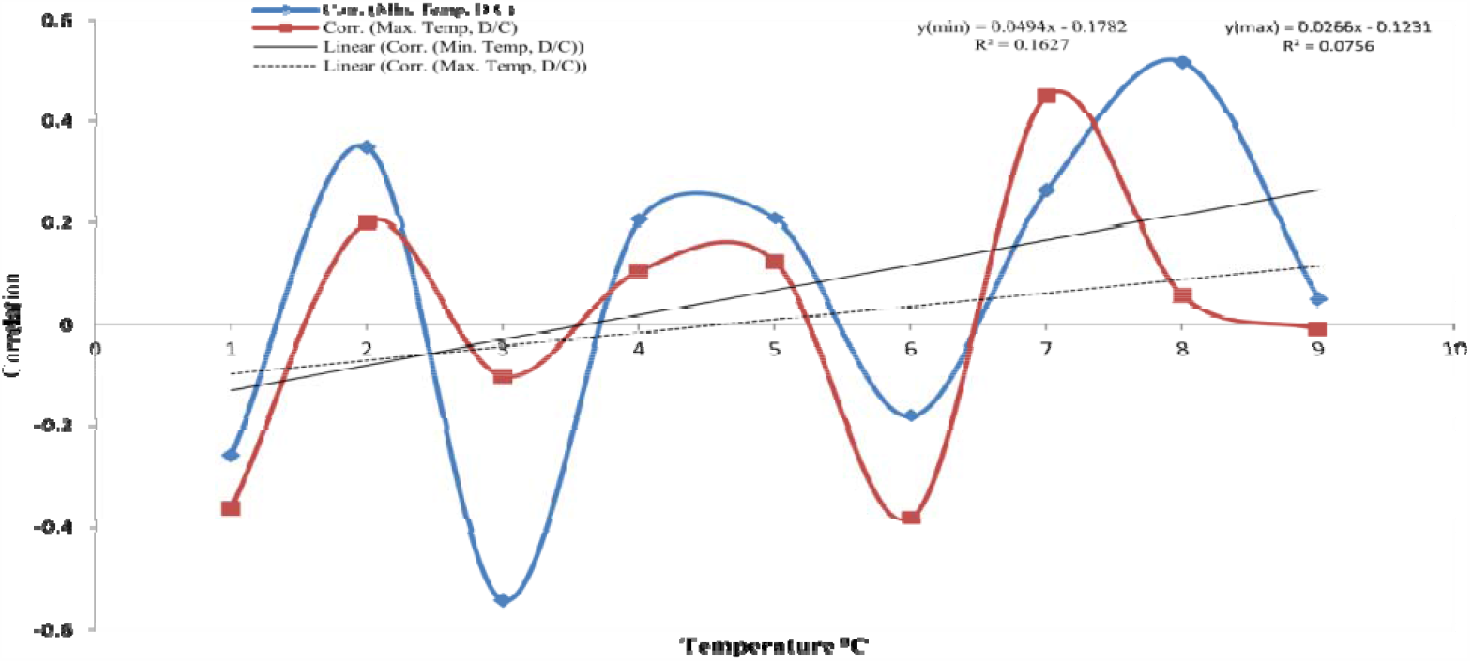
correlation curve between climate temperature and death-to-count (D/C) ratio within the study period.

**Figure 7:**
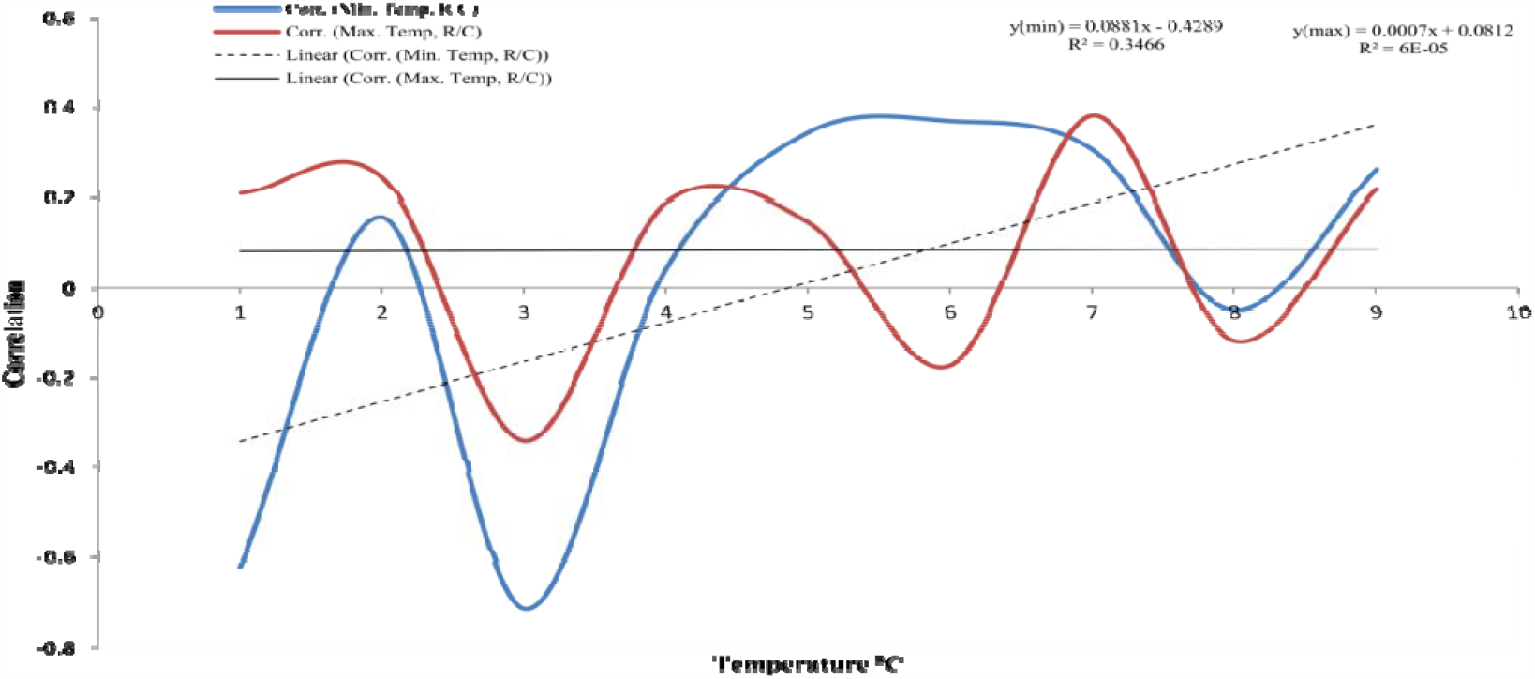
correlation curve between climate temperature and recovery-to-count (R/C) ratio within the study period.

A positive association of r = 0.27, 0.45 for climate temperatures and death-to-count ratio and r = 0.31, 0.38 for climate temperatures and recovery-to-count ratio, respectively, was recorded for the Free State province. In contrast, a negative association of r = -0.54, -0.10 for climate temperatures and death-to-count ratio and r = -0.71, -0.34 for climate temperatures and recovery-to-count ratio, respectively, wa recorded for the Western Cape Province.

Generally, the correlation curve (Figure 6) tends not to show an association between clim temperature and death rates of COVID-19. The curve shows similar trends for both average minimum and maximum climate temperatures. However, the correlation curve (Figure 7) tends to show an association between climate temperature and recovery rates of COVID-19 patients with an almost perfect linear recovery correlation for the maximum temperature of 24°C and above. The curve shows a clear difference in recovery rates at the minimum and maximum climate temperature conditions.

The results show no significant difference in counts, death, and death-to-count rates for different climate temperatures compared with values from other countries with similar and higher climate temperatures, just like South Africa’s. Higher recovery rates for countries with higher climate temperatures (Table 3) have also been reported. This study concludes that the virus can be spread at every climate temperature, with no significant difference in the count and death rates. However, higher recovery rates were recorded for countries with higher climate temperatures than South Africa, which coincides with the results from provinces with higher climate temperatures from the study areas having higher recovery rates of patients infected with COVID-19.

**Table 3:**
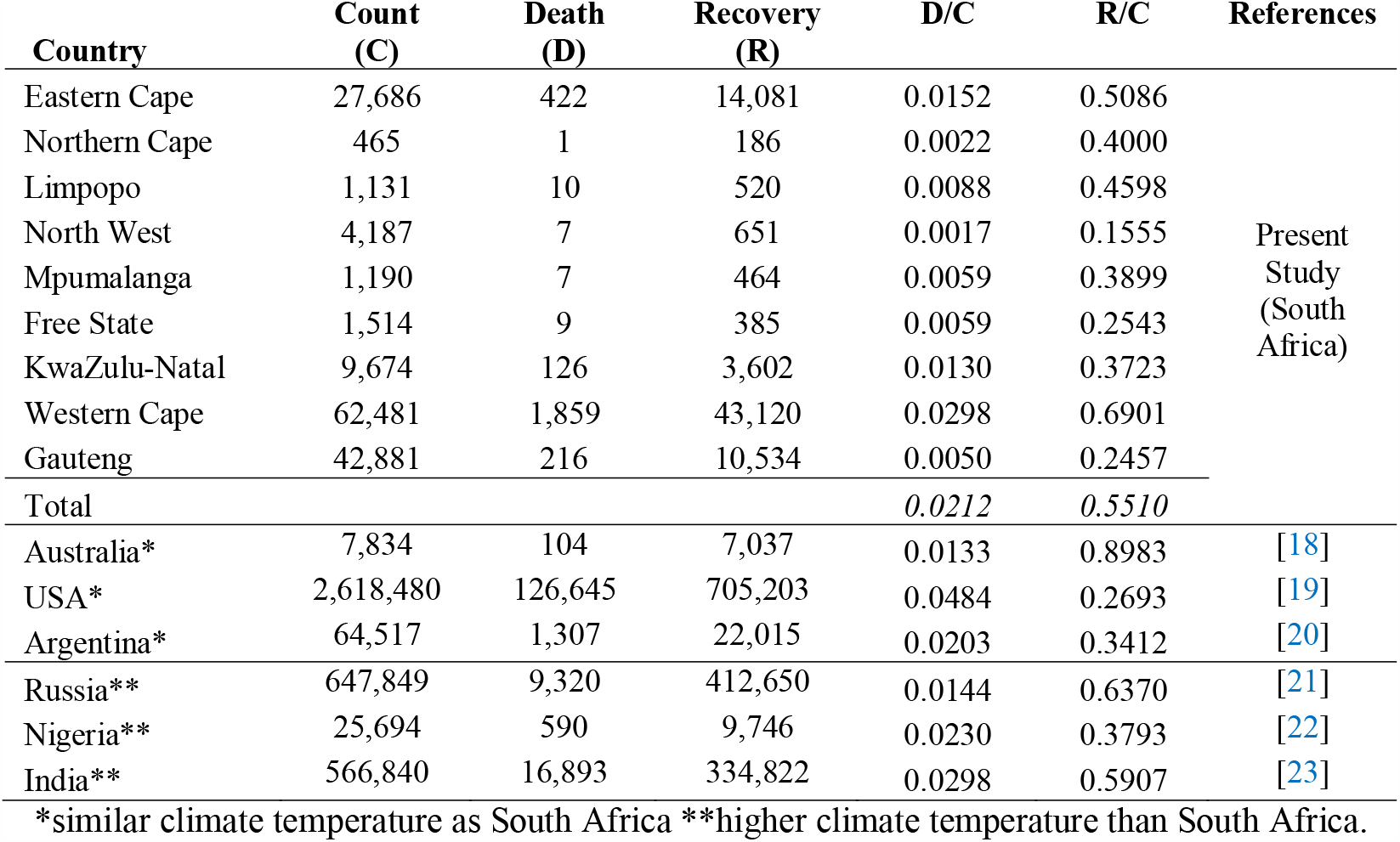
Comparison of COVID-19 counts, death, recovery rates, and death-to-count, recovery-to-count ratios from the study provinces with values from other countries (as of June 30, 2020).

## 4. Conclusion

The study investigates the impact of climate temperature on the counts, recovery, and death rates of COVID-19 cases in all South Africa’s provinces. The findings were compared with those of countries with comparable climate temperatures as South Africa. Our result indicates that a higher or lower climate temperature does not prevent or delay the spread and death rates but shows significant positive impacts on the recovery rates of COVID-19 patients. Warm climate temperatures seem not to restrict the spread of the COVID-19 as the count rate was substantial at every climate temperatures. Thus, it indicates that the climate temperature is unlikely to impose a strict limit on the spread of COVID-19. There is no correlation between the cases and death rates, an indicator that there is no particular temperature range of the climatic conditions closely associated with a faster or slower death rate of COVID-19 patients. However, other shortcomings in this study’s process should not be ignored. Some other factors may have contributed to recovery rates, such as the South African government’s timely intervention to announce a national lockout at the early stage of the outbreak, the availability of intensive medical care, and social distancing effects. Nevertheless, this study shows that a warm climate temperature can only help COVID-19 patients recover more quickly, thereby having huge impacts on the death and active case rates.

## Data Availability

All data in this manuscript were open-sourced from the official reporting agency's websites. https://sacoronavirus.co.za, https://www.weathersa.co.za

## Author contributions statement

NC conceived the study, supervised and guided the process of the research. BA and AOI performed data collection and analysis. All authors wrote and reviewed the manuscript.

